# Association between the COVID-19 pandemic and pertussis in France using multiple nationwide data sources

**DOI:** 10.1101/2021.07.16.21260367

**Authors:** Soraya Matczak, Corinne Levy, Camille Fortas, Jérémie F. Cohen, Stéphane Béchet, Fatima Aït El Belghiti, Sophie Guillot, Sabine Trombert-Paolantoni, Véronique Jacomo, Yann Savitch, Juliette Paireau, Sylvain Brisse, Nicole Guiso, Daniel Lévy-Bruhl, Robert Cohen, Julie Toubiana

**Author notes:** **Corresponding author:** Julie Toubiana, Hôpital Necker – Enfants malades, APHP & Université de Paris, 149 rue de Sèvres, 75015 Paris, France, UE.

## Abstract

**Background:** Interventions to mitigate coronavirus disease 19 (COVID-19) pandemic may impact other respiratory diseases such as pertussis. We aimed to study the course of pertussis in France over an 8-year period and its association with COVID-19 mitigation strategies, using multiple nationwide data sources.

**Methods:** We analyzed the number of French pertussis cases between 2013 and 2020, using the PCR test results from nationwide outpatient laboratories (Source 1) and the pediatric network of 41 hospitals (Source 2), and using the reports of an office-based pediatric national network (Source 3). We conducted a quasi-experimental interrupted time-series analysis, relying on negative binomial regression models. The models accounted for seasonality, long-term cycles, and secular trend, and included a binary variable for the first national lockdown (ordered on March 16, 2021).

**Results:** We identified 19,039 cases of pertussis from the three data sources during the study period. There was a significant decrease of pertussis cases following the implementation of mitigation measures, with adjusted incidence rate ratios of 0.102 (95% CI 0.040-0.256) and 0.216 (95% CI 0.071-0.656) for Source 1 and Source 2, respectively. The association was confirmed in Source 3 (median of 1 [IQR 0-2] vs. 0 [IQR 0-0] pertussis cases per month before and after lockdown, respectively, p=0.0048).

**Conclusion:** The drastic reduction of outpatient and hospitalized cases of pertussis strongly suggests an impact of COVID-19 mitigation measures and their consequences on pertussis epidemiology. Pertussis vaccination recommendations should be carefully followed, and disease monitoring should be continued to detect any resurgence after relaxation of mitigation measures.

**Funding:** There was no specific funding for the study.

## INTRODUCTION

Whooping cough or pertussis is a highly contagious respiratory disease transmitted from human to human via aerosolized respiratory droplets, and is mainly caused by *Bordetella pertussis*. A resurgence of pertussis has been observed in many countries during the last decade despite widespread vaccine implementation.^1^ Most pertussis morbidity and mortality are due to severe clinical forms in young infants that usually require admission to intensive care units. In 2014, the World Health Organization (WHO) estimated pertussis as a cause of 160,700 deaths in children less than five years-old.^2^

Since December 2019, the world has been facing another respiratory infectious disease, the Coronavirus disease 2019 (COVID-19) pandemic due to SARS-CoV-2.^3^ In France, the lockdown was ordered at the beginning of the first wave of the COVID-19 pandemic on March 16, 2020. Measures included school closures, shortly followed by face covering in public places on May 11, 2020.^4^ During the following months, the French government successively introduced several bundles of COVID-19 mitigation measures based upon the dynamics of SARS-CoV-2 infection. The French population initially faced difficulties of access to clinical and biological diagnosis, and delay of childhood vaccinations despite the mandatory infant vaccinations since 2018 and booster vaccination guidelines.^5^ We hypothesized that such measures and their consequences might have impacted pertussis epidemiology, as suggested for other transmissible airborne infectious diseases.^6^

Thanks to a well-established surveillance system of pertussis cases through networks of outpatient laboratories and voluntary hospitals, and the French ambulatory surveillance for outpatient pediatric cases, we assessed the course of pertussis epidemiology in France between January 1, 2013, and December 31, 2020, and its potential association with COVID-19 mitigation strategies.

## METHODS

### Data sources and case definitions

We performed a retrospective analysis using three nationwide data sources from pertussis surveillance systems in France. Those include general population surveillance through two nationwide outpatient laboratories (Cerba [Lab 1] and Biomnis [Lab 2]), which carry out > 90% of the ambulatory testing for pertussis in mainland France (Source 1),^7^ and the monitoring of severe pediatric cases through a nationwide network of 41 voluntary hospitals (Renacoq network, covering about 30% of hospitalized pediatric pertussis cases), collecting hospitalized pertussis cases under 1-year-old (Source 2).^8^ Santé publique France (SpF) and the French National Reference Center (NRC) for *Bordetella* infections collect data every month from Source 1, and twice a year from Source 2. For these two sources, a pertussis case was defined as a positive result for any PCR targeting IS*481* from nasopharyngeal swabs or aspirates. The quantitative PCR methods used for pertussis surveillance were quality-assessed by the French NRC for *Bordetella* infections.^9^

We also included data from the French ambulatory surveillance of whooping cough for outpatient pediatric cases. Implemented in 2002 by the Association Clinique et Thérapeutique Infantile du Val de Marne (ACTIV) and the French NRC for *Bordetella* infections, this surveillance (office-based pediatrician network pertussis surveillance, Source 3) does not aim to estimate the exhaustive number of pertussis cases but to estimate the duration of immunity conferred by pertussis vaccines in children in France.^10^ From 2013 to 2020, 76 pediatricians from the ACTIV network throughout France reported patients with suspected pertussis (cough illness). All suspected patients were invited to perform biological confirmation of pertussis. Cases were defined as either ‘confirmed’ (i.e., positive on culture and/or PCR, or ‘epidemiological’ (i.e., acute cough lasting at least 14 days in a child or adult who was in contact with a confirmed case).^10^

Strengthening the Reporting of Observational studies in Epidemiology (STROBE) guidelines were followed to report the study **(Supplementary Table1**).

### Statistical analysis

We conducted a quasi-experimental interrupted time-series analysis relying on negative binomial regression models,^11^ with a time-unit of one month, to analyze the changes in the incidence of positive *Bordetella* PCRs over time. The models accounted for seasonality (using pairs of sine/cosine terms), secular linear trend, a binary variable to define periods before and after the 1^st^ lockdown (i.e., April 1^st^, 2020, **Supplementary Figure**), and over-dispersion of data. The models also included a dummy variable (with 36-month periods) to adjust for long-term cycles commonly observed in pertussis epidemiology.^8^

First, the analysis was carried out on our most comprehensive data source (Source 1), overall, and then stratified across three pre-specified age groups: pre-school (0-5 years), primary and secondary school children (6-17 years), and adults (≥18 years). We further performed the following sensitivity analyses to assess the robustness of our findings: (1) by fitting the data from Lab 1 and Lab 2 separately, (2) by including the negative samples when available (i.e., modeling the proportion of positive PCR results), (3) by using a different modeling strategy relying on segmented linear regression with autoregressive errors to account for autocorrelation in the data.^11^ Second, the analysis was carried out on data from Source 2, relying on the same negative binomial regression model used for Source 1. Third, the data from Source 3 were not included in the time-series modeling because these data, collected to evaluate the duration of vaccine-induced immunity, did not fulfill model requirements; an analysis comparing the number of pertussis cases per month before and after the lockdown was carried out. Differences between groups were assessed by the Mann-Whitney *U* test or the Student’s *t* test, when appropriate. We used Stata/SE 15.0 (StataCorp LP, College Station, TX, USA) for all analyses.

### Ethics approval

The data collections received approval by Institutional Review and Data protection Board (APHP, N° 2021 0608105701) for Source 1, and by the National Review Boards for Source 2 (CNIL N°449199) and Source 3 (CNIL N°1161396). Source 3 was registered at ClinicalTrials.gov (NCT04318431). All data processing and storage comply with the General Data Protection Regulation (GDPR) and ethical standards of the National Research Committee.

## RESULTS

### Outpatient laboratory pertussis surveillance (Source 1)

Over the study period, there were 17,912 pertussis cases reported from the two private laboratories (Source 1). The median age of patients with a positive PCR was 20.0 (interquartile range [IQR] 6.5-46.2) years, and distribution among age groups was 23% for the 0 – 5 years, 24% for the 6 – 17 years, and 53% for the ≥ 18 years. A subset of cases aged 2 – 20 years old between 2013 and 2019 has already been analysed elsewhere for other purposes.^7^ **Figure 1A** shows the evolution of overall pertussis cases for the 96-month study period. Before the lockdown (before April, 1^st^, 2020), the average number of cases per month was 204 (standard deviation [SD] 119), with a seasonal pattern (i.e., highest incidence in July months, mean 315 [SD 180]). Long-term epidemic cycles were observed: pertussis incidence was higher in 2013 and 2017-2019, compared to 2014-2016. After the lockdown, the average number of cases was much lower, reaching 14 (SD 18) cases per month (p<0.001). Through time-series modeling, we observed a decrease by 89.8% (95% Confidence Interval [CI] 74.4; 96.0%) of pertussis cases after the lockdown; adjusted Incidence Rate Ratio (aIRR) 0.102 (95% CI 0.040; 0.256) (**Table 1)**. This significant decrease was confirmed in all age groups, with a sharper decrease in the 6 – 17 years (−92.7% [95%CI 79.9; 97.3]) and ≥18 years (−93.6% [95% CI 83.5; 97.5]) groups, as compared with the 0 – 5-year group (−78.3% [95% CI 43.6; 91.6], **Table 1**).

**Table 1.** Association between COVID-19 pandemic and pertussis in France: interrupted time-series analysis (2013-2020)

|  |  | Negative binomial modeling |  | Segmented linear regression |  |
| --- | --- | --- | --- | --- | --- |
| Outcome measures |  | Adjusted IRR (95%CI) <sup>1</sup> | p-value | Change in level (95% CI) <sup>2</sup> | p-value |
| **Source 1 – Outpatient laboratories** |  |  |  |  |  |
| Overall number of positive PCRs |  | 0.102 (0.040; 0.256) | <0.001 | - 242.2 (-348.3; -136.2) | <0.001 |
| Number of positive PCRs, by age |  |  |  |  |  |
|  | 0-5 yrs. | 0.217 (0.084; 0.564) | 0.002 | - 45.9 (-83.2; - 8.5) | 0.017 |
|  | 6-17 yrs. | 0.073 (0.027; 0.201) | <0.001 | - 61.2 (-84.5; - 37.9) | <0.001 |
|  | ≥18 yrs. | 0.064 (0.025; 0.165) | <0.001 | -133.6 (-184.2; -83.1) | <0.001 |
| Number of positive PCRs, by Lab |  |  |  |  |  |
|  | Lab1 | 0.093 (0.035; 0.243) | <0.001 | -103.5 (-152.9; -54.1) | <0.001 |
|  | Lab 2 | 0.105 (0.041; 0.265) | <0.001 | -138.7 (-197.3; -80.2) | <0.001 |
| Proportion of positive PCRs* |  | 0.667 (0.016; 27.73) | 0.832 | - 0.04 (-0.125; -0.037) | 0.279 |
| **Source 2 – Hospital laboratories (Renacoq network)** |  |  |  |  |  |
| Number of positive PCRs < 1yr |  | 0.216 (0.071; 0.656) | 0.007 | - | - |

IRR: Incidence Rate Ratio; <sup>1</sup>the estimated IRR was adjusted for long-term cycles, seasonality (using pairs of sine/cosine terms) and secular trend; <sup>2</sup> the estimated change in level was adjusted for long-term cycles, seasonality (adjustment by calendar month), year, and secular trend. \*Proportion calculated using Lab 1 data only for 2013-2015, and then data from Lab 1 and Lab 2 for 2016-2020, because of missing data on negative test results for Lab 2 for 2013-2015.

**Figure 1.**
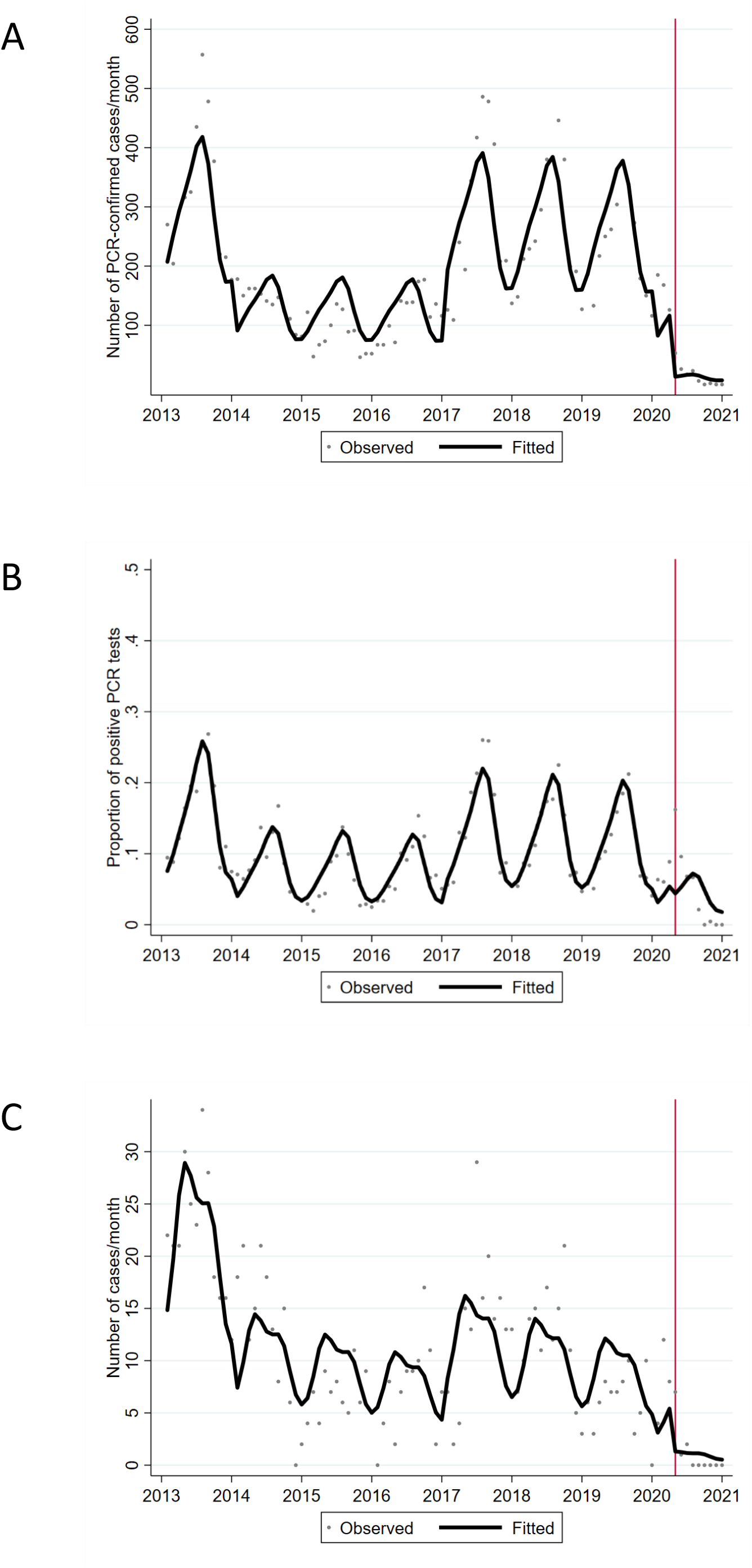
Association between COVID-19 pandemic and pertussis in France: time-series analysis (2013-2020). **A**. Time series of the number of PCR-confirmed cases by month identified by private outpatient laboratories (Source 1). **B**. Proportion of positive cases among PCR tests performed (Source 1). **C**. Time series of the number of cases by month identified by hospital laboratories from the Renacoq Network (Source 2). The observed values are represented in dots; the bold line corresponds to values predicted by the negative binomial regression model; the red vertical line corresponds to the beginning of the 1^st^ lockdown.

In sensitivity analysis, our findings did not differ when data from Lab 1 and Lab 2 were fitted separately (**Table 1**). When including the negative samples, we observed a non-significant decrease in the proportion of positive PCR results (aIRR 0.667 (0.016; 27.73); p= 0.832) (**Figure 1B, Table 1**). Whereas this proportion was 13.4 (SD 7.6) % before lockdown, it fell to 8.3 (SD 5.2) % during the next five months, and to 0.1 (SD 0.2)% the final four months of 2020 (September – December 2020) despite a relatively stable number of tests during the post-lockdown period (mean 334 [SD 60] tests per month, **Table 2**). Segmented linear regression models yielded results similar to negative binomial modeling, with a significant drop in the number of positive PCR results across all age groups (**Table 1**).

**Table 2.** Annual number of pertussis cases diagnosed across data sources (2013-2020)

|  | **Pertussis cases** | **2013** | **2014** | **2015** | **2016** | **2017** | **2018** | **2019** | **2020** |
| --- | --- | --- | --- | --- | --- | --- | --- | --- | --- |
| **Source 1 – IS481 PCR tests from outpatient laboratories** |  |  |  |  |  |  |  |  |  |
| **Lab 1** | Positive | 2,266 | 962 | 450 | 655 | 1,473 | 1,369 | 1,095 | 206 |
|  | Negative | 13,139 | 11,216 | 8,614 | 9,112 | 10,355 | 12,623 | 11,336 | 4,152 |
|  | Total number of tests | 15,405 | 12,178 | 9,064 | 9,767 | 11,828 | 13,992 | 12,431 | 4,358 |
| **Lab 2** | Positive | 1,601 | 666 | 552 | 784 | 1,881 | 1,864 | 1,688 | 400 |
|  | Negative | N/A | N/A | N/A | 9,372 | 11,296 | 13,069 | 13,819 | 5,366 |
|  | Total number of tests | 1,601 | 666 | 552 | 10,156 | 13,177 | 14,933 | 15,507 | 5,766 |
| **Source 2 – Hospital laboratories (Renacoq network)** |  |  |  |  |  |  |  |  |  |
|  | Number of positive tests | 266 | 149 | 81 | 86 | 162 | 141 | 73 | 34 |
| **Source 3 – Outpatient pediatric network (ACTIV)** |  |  |  |  |  |  |  |  |  |
|  | Total clinically suspected pertussis | 48 | 27 | 16 | 15 | 37 | 32 | 38 | 7 |
|  | ‘Confirmed’ cases | 14 | 13 | 4 | 5 | 14 | 10 | 14 | 2 |
|  | ‘Epidemiological’ cases | 7 | 4 | 4 | 4 | 13 | 6 | 17 | 4 |

N/A: not available.

### Hospital-based pertussis surveillance (Source 2)

Over the study period, 992 positive PCR tests were collected through the Renacoq hospital network (Source 2). Distribution among age groups was 61% for the 0 – 2 months, 25% for the 3-5 months, and 14% for the 6-11 months. We also observed a seasonal pattern (**Figure 1C)**. Before lockdown, the average number of pertussis cases per month was 11.3 (SD 7.3). After the lockdown, a sharp decrease in the number of hospitalized pertussis cases was confirmed, with an average number of the case reaching 1.1 (SD 2.3), p<0.001. In time-series modeling, there was a significant association between COVID-19 mitigation measures and pertussis cases, with a reduction of 78.4% (95%CI 34.4; 92.9); aIRR 0.216 (95% CI 0.071; 0.656); p=0.007 (**Figure 1C, Table 1 and 2**). Between July 2020 and December 2020, no case of pertussis was notified.

### Office-based pediatrician network pertussis surveillance (Source 3)

From 2013 to 2020, the pediatric outpatient network (Source 3) reported 135 cases of pertussis. Confirmed and epidemiological cases accounted for 76 (56.3%) and 59 (43.7%) cases, respectively (**Table 2**). Among the 59 epidemiological cases, 25 were adults (family members), and 34 were children (siblings, family members, daycare center, or school). The mean age of confirmed cases was 7.1 (SD 4.2) years and of epidemiological cases was 22.4 (SD 20.1) years. Before the lockdown, the median number of cases per month was 1 (interquartile range [IQR] 0-2). After the lockdown, no case of pertussis was identified in Source 3 (median 0, IQR 0-0; p=0.0048).

## DISCUSSION

In this 8-year population-based study, we described the dynamics of pertussis epidemiology in France before and following the implementation of the first COVID-19 mitigation measures (March 16, 2020, school closure and national lockdown, **Supplementary Figure1**) ^4^. All our data sources highlighted a long-term cyclical pattern commonly observed in pertussis ^8^, with two high-incidence periods: the year 2013, corresponding to the 2011-2013 epidemic observed in many countries ^7,12^, and years 2017-2019. Therefore, a decrease in pertussis incidence was to be expected in 2020, such as for the 2014-2016 period. However, our interrupted time-series analysis, accounting for this 36-month pertussis cycle and seasonality, showed an even sharper decrease in pertussis positive PCR tests, with a strong evidence of its association with anti-COVID-19 measures. This drop was particularly marked, with a number of pertussis cases close to null after the lockdown, a level never observed since the beginning of the French pertussis surveillance in 1996 ^7^.

All age groups were affected by the decrease of pertussis cases following the mitigation measures. However, a lower decrease was observed in the 0-5-year group tested by the two private laboratories. One explanation could be that the youngest children were more likely to visit their pediatrician despite lockdown measures. Another explanation could be that transmission might still have been active in this age group during the first months of 2020. We did not find a significant association between the proportion of positive PCRs and the mitigation measures against SARS-CoV-2. Our data show that the overall number of PCR tests decreased right after the lockdown, while pertussis might still have circulated in households. However, despite a steady number of PCR tests performed during the whole post-lockdown period, the number of positive cases reached almost zero within the last 4 months of 2020; of note access to clinical and biological diagnosis went back to normal during that period. All these findings suggest an important decrease of pertussis circulation in France.

Previous studies revealed a major impact of COVID-19 pandemic mitigation measures on transmissible diseases, such as, in the pediatric population, gastroenteritis, common cold, bronchiolitis, and acute otitis media.^6^ However, a resurgence of some other airborne infectious diseases has been observed in the setting of relaxed physical distancing recommendations, such as the delayed inter-seasonal Respiratory Syncytial Virus (RSV) epidemic described by Foley *et al*.^13^ For now, we did not observe any pertussis resurgence despite the end of the first lockdown and school fully reopening in France. Our data suggest that, unlike RSV, slight relaxation of public health measures (with maintained social distancing and mask-wearing) has no impact on pertussis dynamics, as for the influenza virus for which no outbreak has been identified worldwide this year.^14,15^ We identified five studies from Australia, South Korea, and Italy, describing the trends of several infectious diseases, including pertussis, before and after implementing their respective COVID-19 mitigation measures.^16-19^ Although the power of these studies was much lower compared to ours (lower duration of surveillance and description of a maximum of ∼1500 cases, vs. 19,039 cases in our study), they all found a decrease in pertussis cases after the implementation of mitigation measures, with a reduction ranging from 28% to 87%.^16-20^

Even if the incidence of pertussis has strongly decreased since the introduction of booster vaccines, *B. pertussis* is still circulating in France, especially in older children, adults and the elderly, as we have previously shown.^7,8,10^ This may be due to non-optimal vaccination coverage in these populations,^21^ and waning of immunity with the acellular pertussis vaccine.^7,22^ The limited circulation of *B. pertussis* in the community due to the measures imposed by the COVID-19 pandemic is a clear positive collateral effect. However, this reduced bacterial circulation, raises concern about a potential decrease in herd immunity and the risk of a larger epidemic in the coming years. During the lockdown, pediatricians involved in data Source 3 declared following vaccinations guidelines.^23^ However, as in many countries, delays in vaccination were observed in France in the general population since the beginning of the pandemic; they mostly concerned booster vaccinations for children and adults.^5,24^ Such delays can be dangerous for infants under 6 months of age who are non- or partially vaccinated, notably if there is no catch-up before a potential resurgence of pertussis. Indeed, as many other community-acquired infections, the dramatic decrease of pertussis incidence could suggest an epidemic rebound after relaxation of all mitigation measures.^25^ Therefore, vaccination guidelines (infants, children, adolescents, adults and cocooning strategy) should be strictly followed to increase herd immunity, and maternal vaccination against pertussis during pregnancy, not yet implemented in France, should presently be considered by French national health institutions.^26^

Our study has limitations. First, measurement bias is present in Sources 1 and 2 as our pertussis case definition only considered positive PCRs. We therefore might have missed cases diagnosed by serology (sometimes prescribed even if not recommended), or based on clinical grounds, especially during the 1^st^ lockdown (from mid-March to May, 2020), when transportation was limited, office-based physicians unreachable, and private laboratories overwhelmed by the implementation of large-scale SARS-CoV-2 testing. Second, we cannot determine whether the decrease of pertussis cases observed here was due to decreased pertussis circulation or reduced testing. However, the similar decrease observed in hospitalized cases in the youngest population does not favor the latter hypothesis, and access retriction to outpatient testing was mostly limited to the first lockdown period. Third, we may have lacked statistical power to detect a significant decrease in the proportion of positive cases, as the post-lockdown period was relatively short compared to the pre-lockdown period (i.e., 9 months vs. 87 months, respectively), and negative PCR tests results were not available from data Source 2.

In conclusion, this national-level study shows a strong association between the COVID-19 pandemic and pertussis in France, with an unprecedented drop of pertussis cases. Pertussis should be closely monitored to detect any resurgence in the community when social distancing restrictions will be relaxed, as well as compliance to vaccination of infants, children, and adults.

## ADDITIONAL INFORMATION

### Contributors

Study conception: JT, CL, RC, DLB, NG, JFC, SBr. Data collection: SM, CF, CL, SBe, SG, FAEB, YS, JP, STP, VJ, JT. Statistical analysis: JFC. Data analysis: JFC, SM, CF, SBe, CL JT. Data interpretation: SM, JT, CL, JC, CF, SBe, SG, FAEB, YS, JP, DLB, NG, SBr, RC. Microbiology analyses: STP, VJ. Drafting the manuscript: SM, JFC, JT. Revising the manuscript: all authors. Approving the final version submitted: all authors. Study supervision: JT.

### Data sharing

Anonymised database will be made available on reasonable request.

### Authors license statement

This research was funded, in whole or in part, by Institut Pasteur and Santé publique France. For the purpose of open access, the authors have applied a CC-BY public copyright license to any Author Manuscript version arising from this submission.

## Acknowledgments

We thank the microbiologists from the Renacoq network, as well as Annie Landier and Nathalie Armatys from the NRC, for their participation to PCR testings. We thank Martin Chalumeau for his methodological advices, and all the pediatricians of the French ambulatory surveillance of whooping cough. We thank Isabelle Ramay, Karin Lejeune, Aurore Prieur, and Marine Borg for the technical assistance

