## Supplementary Figure for "Association between the COVID-19 pandemic and pertussis in France using multiple nationwide data sources"

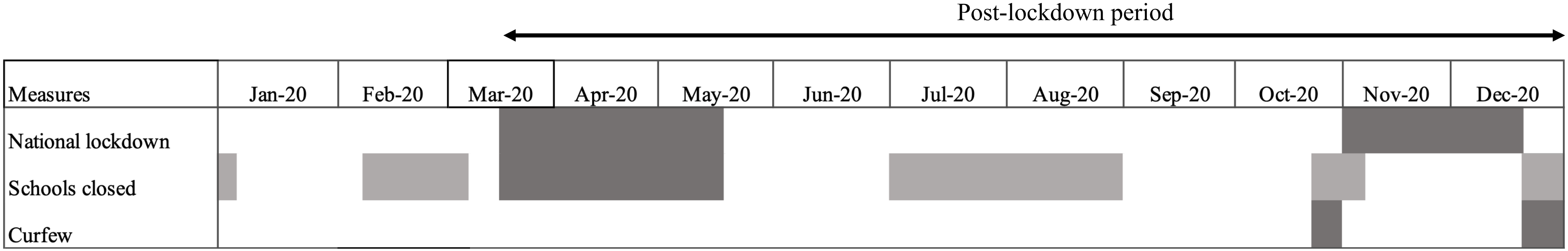


**Supplementary Figure 1. French COVID-19 mitigation measures in 2020.** Mitigation measures are represented in dark gray, periods when schools were closed because of holidays are represented in light gray
